# Against Empathy: distinct correlates of empathy and compassion with burnout and affective symptoms in health professionals and students

**DOI:** 10.1101/19005231

**Authors:** Aline Romani-Sponchiado, Matthew R. Jordan, Argyris Stringaris, Giovanni Abrahão Salum

## Abstract

Concern for the well-being of medical professionals has increased considering the high rates of depression and suicidal ideation observed among medical students and residents. However, the causes of such psychological distress among health professionals are still unknown.
One possibility is that such negative outcomes arise from individual differences in how clinicians respond to the emotional states of their patients: while some tend respond with empathy (feeling what others feel), others tend respond with compassion (caring about what others feel). The aim of this study is to investigate the hypothesis that empathy is related to higher levels of burnout and affective symptoms, while compassion is related to lower levels of these outcomes.
We surveyed 464 undergraduate students and professionals in medicine (34.3%), psychology (47%) and nursing (18.8%), 79.7% female, with a median age of 23.3. The survey included the concern and perspective taking subscales from the Interpersonal Reactivity Index (IRI); empathy and behavioral contagion from the Empathy Index (EI); the depression, anxiety, and anger subscales from PROMIS; and the Medical Student Well-Being Index (MSWBI).
Empathy was associated with higher symptoms of burnout, depression, anxiety and anger; while higher levels of compassion were associated with lower levels of these outcomes.
Our findings provide new evidence that the well-being of medical professionals might be affected differently depending on socio-emotional traits relevant to emotional connection.

---

A recent perspective published in *The Lancet* emphasizes the historical and modern occupational strains of medical professionals^1^. As concern for the well-being of medical professionals has increased, the high rates of depression and suicidal ideation observed among medical students^2^ and residents^3^ have garnered increasing attention. However, the causes of such psychological distress among health professionals are still unknown. One possibility is that such negative outcomes arise from individual differences in how clinicians respond to the emotional states of their patients. Prior work has shown that people differ in how they respond to those who are in distress: while some tend respond with empathy (feeling what others feel), others tend respond with compassion (caring about what others feel)^4^. Here we investigate the hypothesis that empathy is related to higher levels of burnout and affective symptoms, while compassion is related to lower levels of these outcomes.

We surveyed 464 undergraduate students and professionals in medicine (34·3%), psychology (47%) and nursing (18·8%), 79.7% female, with a median age of 23·3. To assess compassion, we used the concern and perspective taking subscales from the Interpersonal Reactivity Index (IRI). To assess empathy, we used the empathy and behavioral contagion from the Empathy Index (EI)^4^. Symptoms of negative affect were assessed using the depression, anxiety, and anger subscales from PROMIS^5^, and burnout was assessed using the Medical Student Well-Being Index (MSWBI). This study was approved by the Ethics Committee from UFRGS.

Empathy was associated with higher symptoms of burnout (β= ·691, p< ·001), depression (β= ·456, p< ·001), anxiety (β= ·669, p< ·001) and anger (β= ·59, p< ·001); higher levels of compassion were associated with lower burnout (β=- ·457, p=·002), depression (β=- ·47, p< ·001), anxiety (β=- ·487, p=·002) and anger (β=- ·642, p< ·001). Post-hoc analysis revealed that results were driven by the Perspective Taking subscale of the IRI and Empathy subscale of the EI (Figure 1).

**Figure 1.**
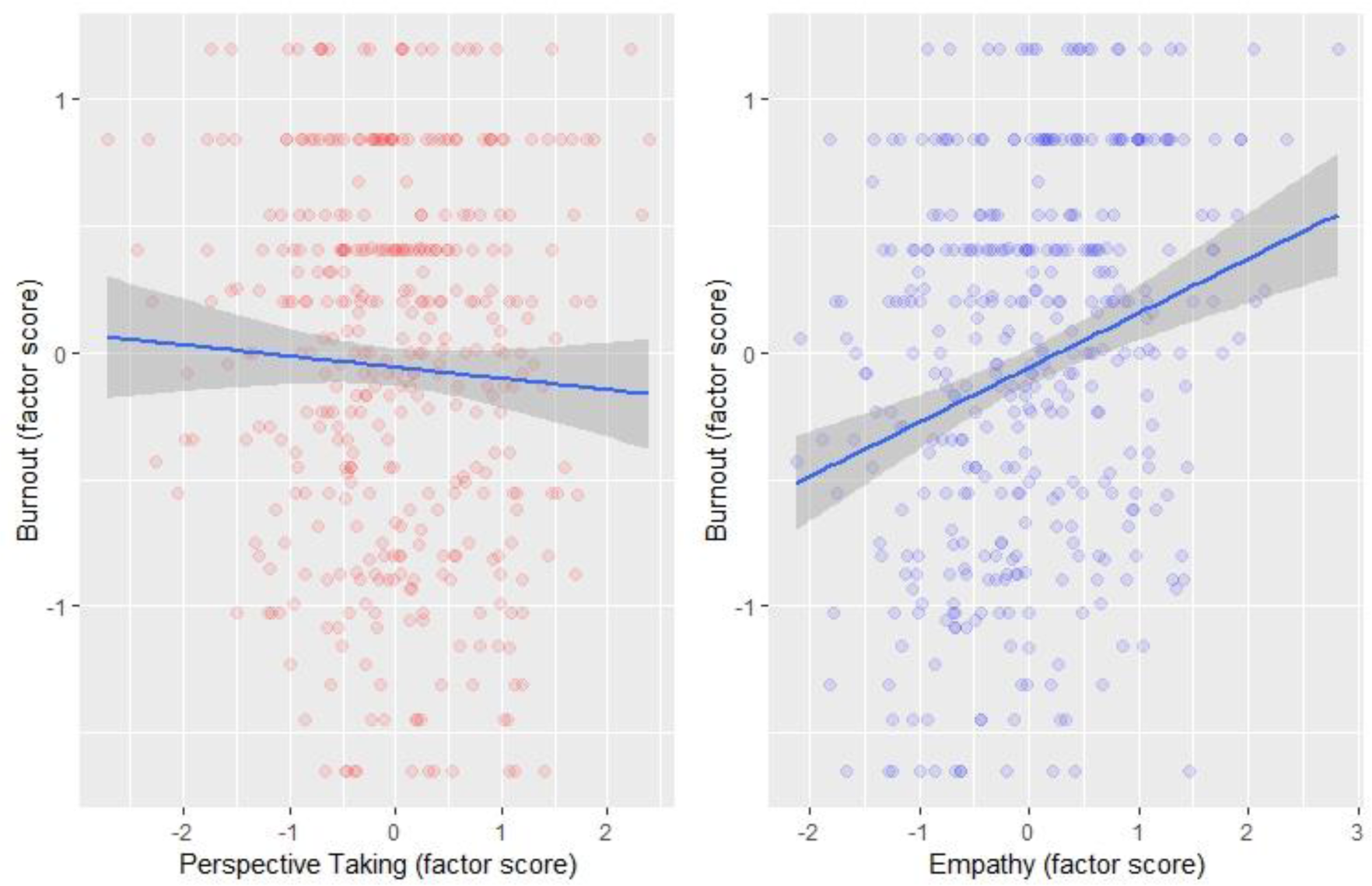
Associations between Perspective Taking and Empathy with burnout Post-hoc analysis showed association of Perspective Taking with lower depression (β= - ·298, p< ·001), anxiety (β= -·313, p< ·001), anger (β= -·415, p< ·001) and burnout (β= - ·267, p=·005). Empathy was associated with higher levels of depression (β= ·33, p=·016), anxiety (β= ·514, p=·001), anger (β= ·373, p=.008) and burnout (β= ·578, p=·006).

Our findings provide new evidence that the well-being of medical professionals might be affected differently depending on socio-emotional traits relevant to emotional connection. These results raise two important issues. First, the medical literature omits the conceptual and psychological distinction between empathy and compassion as two different traits with different socio-emotional effects. Second, to the extent that medical professionals are trained to feel more empathy, such training might have unwanted consequences on their mental health. In contrast, a focus on increasing compassion among medical professionals might have desirable consequences on their well-being.

## Data Availability

All data referred to in the manuscript are available.

